# MODELLING SEASONAL TRENDS OF MALARIA INCIDENCE IN NASARAWA STATE, NIGERIA USING HEALTH FACILITY SURVEILLANCE DATA

**DOI:** 10.64898/2026.05.12.26353062

**Authors:** Grace Iberi Iheanacho, Maxwell Azubike Ijomah, Ibidabo David Alabere

## Abstract

Malaria transmission in Nigeria is highly seasonal and climate-sensitive, yet routine surveillance and meteorological datasets remain underutilized for predictive modelling at subnational levels. This study modelled seasonal malaria incidence trends in Nasarawa State, Nigeria using routine surveillance and climatic data.

A retrospective ecological time-series study was conducted using monthly confirmed malaria incidence data from all 13 Local Government Areas of Nasarawa State between 2021 and 2025. Rainfall and temperature were examined as the climatic predictors. Seasonal decomposition and cross-correlation analyses were performed to identify the temporal patterns and lag structures. Seasonal Autoregressive Integrated Moving Average (SARIMA) and Seasonal Autoregressive Integrated Moving Average with Exogenous Variables (SARIMAX) models were developed using the Box–Jenkins framework. Model performance was evaluated using the Root Mean Square Error (RMSE) and Mean Absolute Percentage Error (MAPE).

Malaria incidence showed pronounced seasonal peaks, with the highest transmission occurring during the rainy season. Cross-correlation analysis identified rainfall at a one-month lag and contemporaneous temperature as significant predictors of malaria incidence. The SARIMAX model outperformed the univariate SARIMA model, achieving strong predictive accuracy (MAPE = 8.7%). Forecast projections indicate sustained transmission with a peak incidence expected between June and August 2026.

Malaria transmission in Nasarawa follows a predictable seasonal pattern that is influenced by climatic variability. Incorporating rainfall and temperature into SARIMAX models improves the forecasting performance and provides evidence supporting climate-informed malaria surveillance and preparedness in endemic settings.

**PLAIN LANGUAGE SUMMARY:** Malaria transmission in Nigeria changes with the seasons and is influenced by weather conditions such as rainfall and temperature. This study analysed malaria surveillance data from Nasarawa State between 2021 and 2025, together with climate data, to identify seasonal patterns and predict future malaria trends. The findings showed that the number of malaria cases increased during the rainy season, particularly between June and August. A forecasting model that includes climate variables predicts malaria trends more accurately than models without climate information. These findings suggest that combining routine surveillance and climate data may improve malaria preparedness and planning in endemic areas.

## INTRODUCTION

Malaria remains a major global public health problem, particularly in sub-Saharan Africa, where climatic and ecological conditions favour sustained transmission [1]. According to the World Health Organization, an estimated 249 million malaria cases and 608,000 malaria-related deaths will occur globally in 2022, with the African region accounting for approximately 94% of all cases and deaths [1]. Nigeria bears the highest malaria burden globally despite the sustained implementation of preventive and control interventions, such as insecticide-treated nets, seasonal malaria chemoprevention, indoor residual spraying, and artemisinin-based combination therapy [1].

Malaria transmission is strongly influenced by climatic and environmental factors, particularly rainfall and temperature, which affect mosquito breeding, vector survival, biting behaviour, and parasite development [2,3]. Rainfall contributes to the formation of stagnant water bodies that serve as breeding habitats for *Anopheles* mosquitoes, whereas temperature influences the rate of parasite maturation within the mosquito vector and overall mosquito longevity [3]. Consequently, the incidence of malaria in endemic tropical settings often demonstrates pronounced seasonal variation.

Several studies have demonstrated that incorporating climatic variables into predictive models improves their ability to forecast malaria outbreaks and seasonal transmission dynamics [4,5]. Time-series forecasting approaches such as Seasonal Autoregressive Integrated Moving Average (SARIMA) models are widely used in infectious disease epidemiology owing to their ability to model temporal autocorrelation, trends, and seasonality [6].

Recent studies in malaria-endemic African settings have further highlighted the growing importance of climate-informed malaria forecasting. Bakare et al. demonstrated the usefulness of SARIMA-based approaches for modelling malaria transmission trends across selected Nigerian states [7]. Similarly, a forecasting study conducted in Sierra Leone reported that integrating climatic variability into Seasonal Autoregressive Integrated Moving Average (SARIMA) and Seasonal Autoregressive Integrated Moving Average with Exogenous Variables (SARIMAX) frameworks improved the prediction accuracy of malaria transmission trends [8]. In Ethiopia, Gelaw and Abera demonstrated improved forecasting performance when rainfall and temperature were incorporated as exogenous regressors into a SARIMAX model for malaria prediction in a resource-limited urban setting [9]. In South Africa, SARIMA-based forecasting approaches have also been applied to support malaria surveillance and elimination planning in the Limpopo Province [10].

Beyond conventional statistical forecasting models, emerging machine learning and deep learning approaches are being increasingly explored for malaria prediction. A recent climate-informed forecasting study using transformer-based deep-learning models demonstrated the utility of high-resolution environmental datasets for malaria prediction [11]. Additionally, generalized linear modelling approaches have been successfully applied for malaria forecasting in endemic regions of Senegal [12]. Despite these advances, the SARIMA and SARIMAX models remain particularly valuable in low-resource settings, because they are interpretable, computationally efficient, and compatible with routinely collected surveillance data.

In Nigeria, routine malaria surveillance data are systematically collected through the District Health Information Software 2 (DHIS2) platform, while meteorological data are available through the Nigerian Meteorological Agency (NiMet). However, these datasets are predominantly used for retrospective reporting and program monitoring rather than predictive modelling and forecasting, particularly at the subnational level [13].

Nasarawa State provides an appropriate setting for climate-sensitive malaria modelling because of its endemic malaria profile, tropical savannah climate, and marked wet and dry seasonal patterns. Annual rainfall patterns and temperature variability may substantially influence vector ecology and malaria transmission dynamics in the state [14]. Furthermore, relatively consistent health facility reporting through the DHIS2 provides an opportunity to utilize routine surveillance data for predictive analytics.

Seasonal Autoregressive Integrated Moving Average models incorporating exogenous climatic variables (SARIMAX) provide a robust framework for modelling seasonal malaria transmission while accounting for environmental predictors. Integrating lagged climatic variables into malaria forecasting models may improve the predictive performance and strengthen climate-informed malaria surveillance systems.

Therefore, this study aimed to model and forecast seasonal trends in malaria incidence in Nasarawa using routine health facility surveillance data and climatic variables.

## MATERIALS AND METHOD

### Study Design

This retrospective ecological time-series study was conducted using routinely collected malaria surveillance and meteorological data from January 2021 to December 2025.

### Study Area

This study was conducted in Nasarawa State, North-Central Nigeria. The state experiences a tropical climate characterized by distinct wet and dry seasons, annual rainfall ranging from 1,000 mm to 1,750 mm, and mean temperatures between 25°C and 34°C. The ecology of the state supports mosquito breeding and sustains malaria transmission.

### Data Sources

Monthly confirmed malaria case data were obtained from the District Health Information Software 2 (DHIS2) platform across all 13 Local Government Areas (LGAs) of Nasarawa State. Climatic data, including monthly rainfall and mean temperature, were obtained from the Nigerian Meteorological Agency (NiMet). The population estimates used for incidence calculations were obtained from the National Population Commission.

### Study Variables

The primary outcome variable was monthly malaria incidence per 1,000 individuals. The explanatory variables included monthly rainfall and mean monthly temperature.

### Data Processing and Time-Series Preparation

The monthly malaria case counts were aggregated across all LGAs to generate a state-level time series. Malaria incidence rates were calculated using monthly confirmed malaria cases divided by projected population estimates and were expressed per 1,000 population.

Climatic variables were synchronized temporally with the malaria incidence data prior to the analysis. Missing observations were assessed and no major data discontinuities requiring imputation were identified.

### Statistical Analysis

Descriptive analyses were conducted to summarise malaria incidence and climatic variability over time.

Time-series decomposition was performed to evaluate the trend and seasonal and residual components of malaria incidence. Seasonal decomposition plots were generated to assess the temporal structure and recurring transmission patterns.

The stationarity of the malaria incidence series was assessed using visual inspection, autocorrelation function (ACF) plots, partial autocorrelation function (PACF) plots, and differencing procedures where necessary.

Cross-correlation function (CCF) analysis was used to identify lag relationships between the climatic variables and malaria incidence. The lag structures were evaluated to determine the delayed effects of rainfall and temperature on malaria transmission.

Three model classes were evaluated sequentially:

1. Autoregressive Integrated Moving Average (ARIMA)
2. Seasonal Autoregressive Integrated Moving Average (SARIMA)
3. Seasonal Autoregressive Integrated Moving Average with Exogenous Variables (SARIMAX)

The model development followed the Box–Jenkins methodology [6]. Candidate models were selected based on the AIC values, residual diagnostics, forecasting performance, and epidemiological plausibility.

Model performance was evaluated using:

- Root Mean Square Error (RMSE)
- Mean Absolute Error (MAE)
- Mean Absolute Percentage Error (MAPE)

Forecast validation was conducted using holdout validation procedures, in which data from 2025 were used as the test dataset.

Residual diagnostics, including residual plots, autocorrelation analysis, and distributional assessment, were performed to assess model adequacy.

Statistical analyses were conducted using Python version XX and relevant time-series modelling libraries.

### Ethical Considerations

Ethical approval and administrative permission were obtained from relevant authorities prior to data collection and analysis. Only aggregated secondary data were used and no patient identifiers were included.

## RESULTS

### Seasonal Trends of Malaria Incidence

The monthly malaria incidence demonstrated marked seasonal variation over the study period (2021–2025), with recurrent peaks occurring during the rainy season.

Time-series analysis revealed increased malaria transmission between June and September, with the peak incidence typically occurring in August (Figure 1).

**Figure 1:**
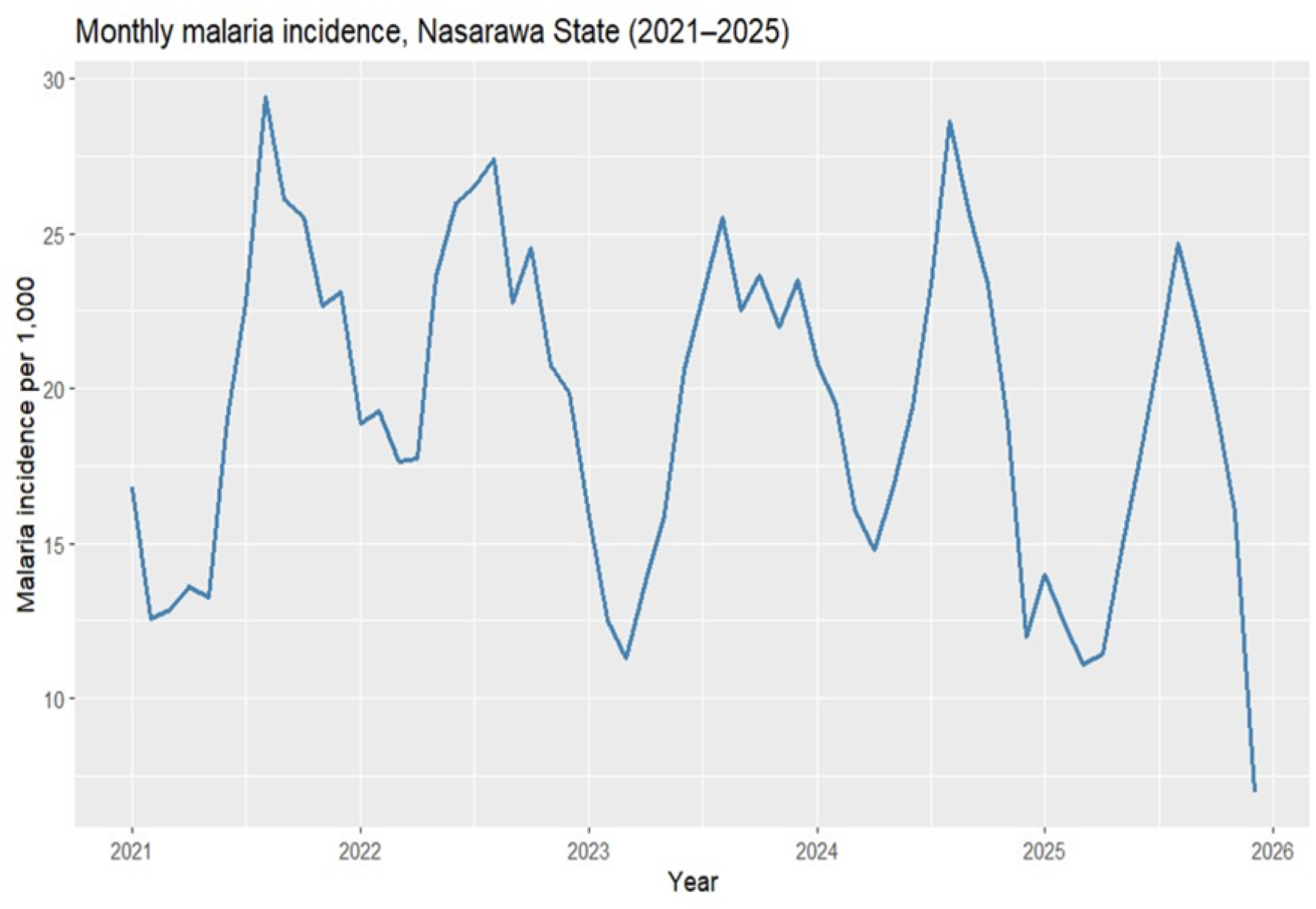
Monthly Malaria incidence timeseries in Nasarawa State 2021-2025. Malaria incidence expressed as cases per 1,000 population per month. Data represent aggregated confirmed malaria cases across 13 Local Government Areas (LGAs) in Nasarawa State. Peaks in incidence correspond to rainy season months; troughs represent dry season periods.

The seasonal decomposition of the time series confirmed a strong seasonal component superimposed on a relatively stable long-term trend (Figure 2). No major structural anomalies were observed in the residual components.

**Figure 2:**
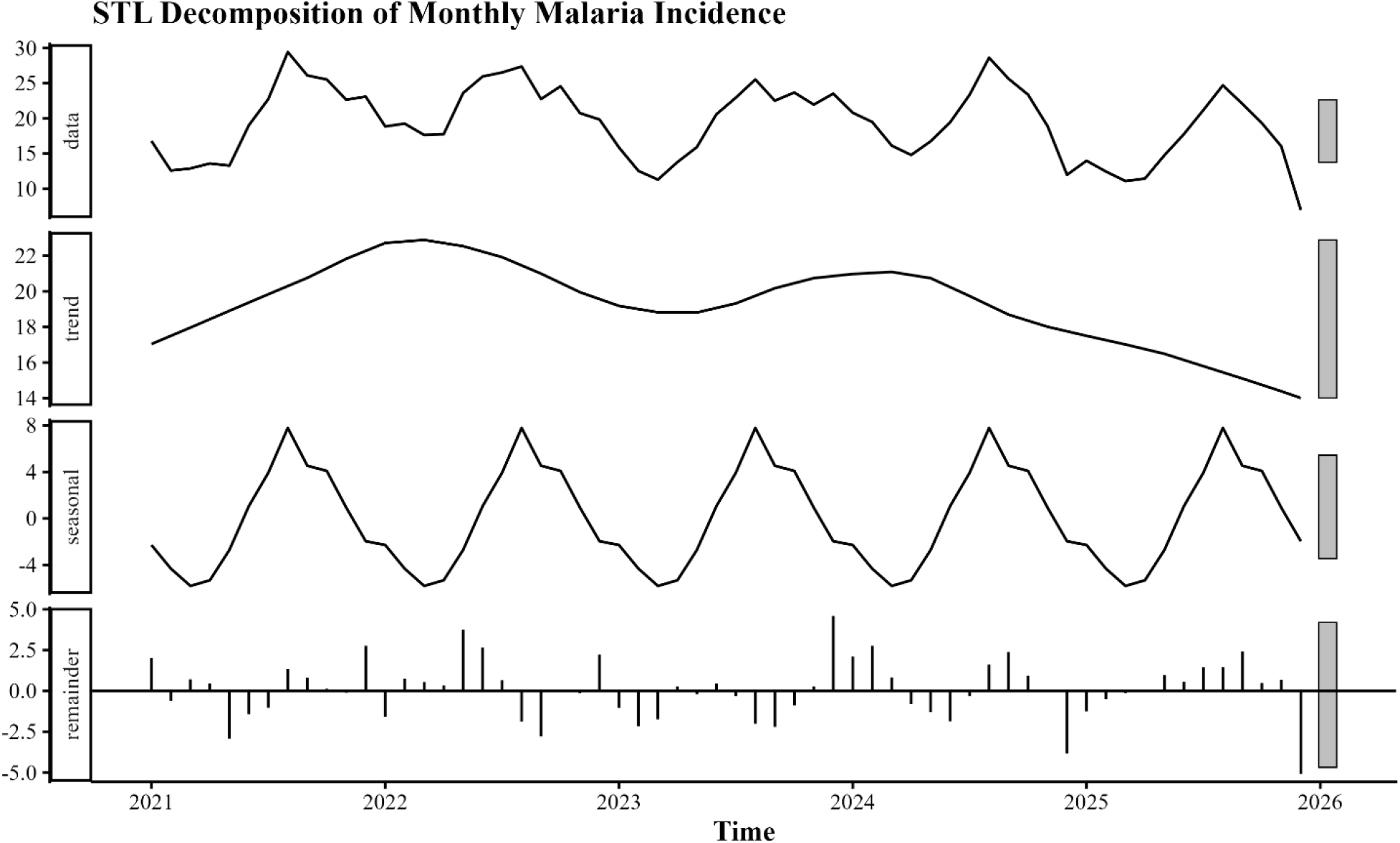
Seasonal decomposition of monthly malaria incidence in Nasarawa State. Trend, seasonal, and residual components derived from time-series decomposition of monthly malaria incidence between 2021 and 2025.

### Climatic Correlates of malaria incidence

Cross-correlation analysis identified rainfall at a one-month lag as positively associated with malaria incidence, while contemporaneous temperature demonstrated a significant inverse relationship with malaria transmission (Figure 3).

**Figure 3:**
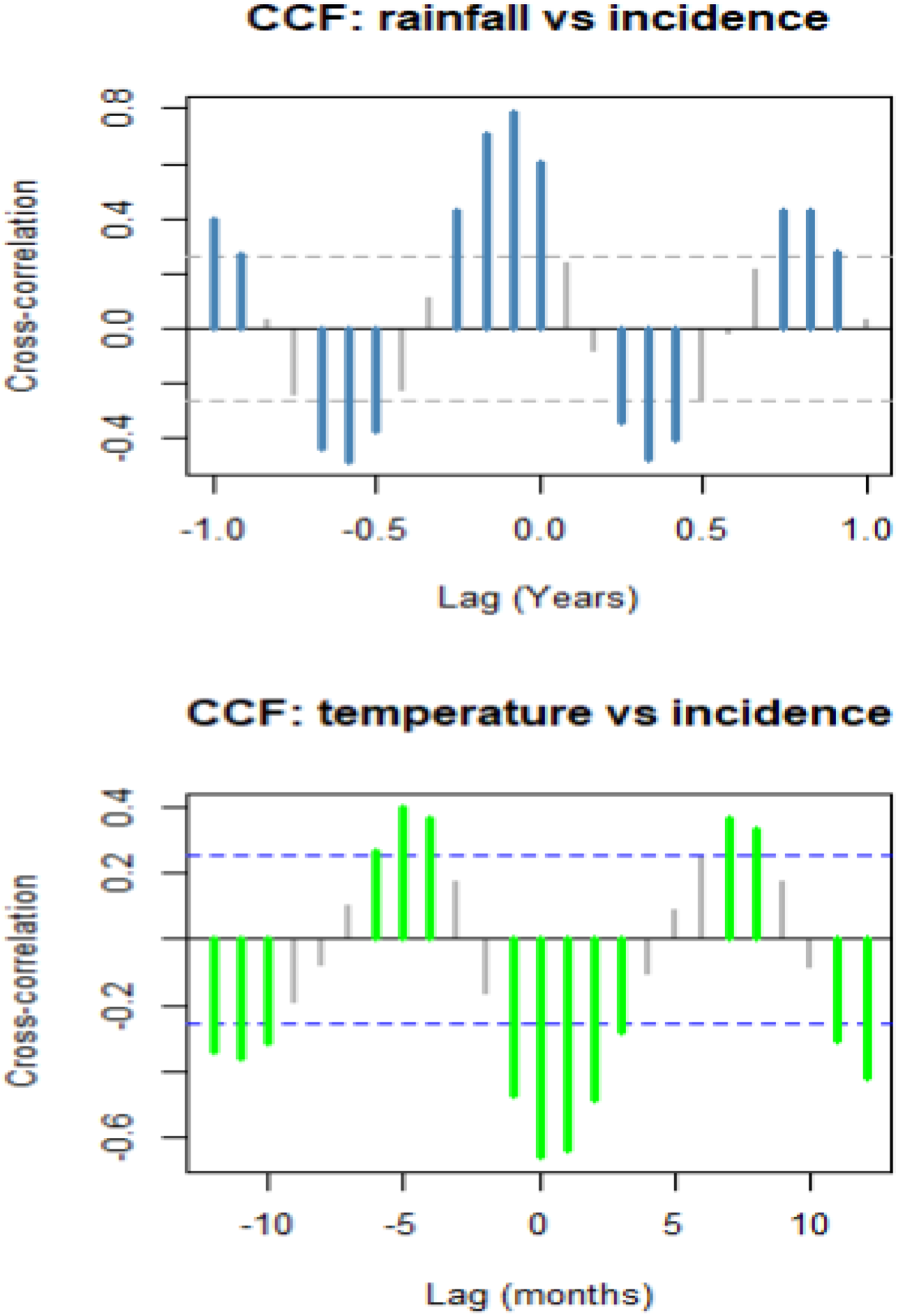
Cross correlation between climatic variables with malaria cases at different lag periods. Cross-correlation function (CCF) plots showing lag relationships between rainfall, temperature, and malaria incidence

These lag structures were consistent across most LGAs with minimal spatial heterogeneity in the lag direction. The summary of the correlation and lag estimates are listed in Table 1.

**Table 1:** Estimated lag relationships between climatic variables and malaria incidence.

| Climate Variable | Optimal Lag (Months) | CCF Value | Direction of Association |
| --- | --- | --- | --- |
| Rainfall | 1 | Positive | Increased rainfall associated with increased malaria incidence after one month |
| Temperature | 0 | Negative | Increased temperature associated with reduced malaria incidence contemporaneously |

**Table 1:**
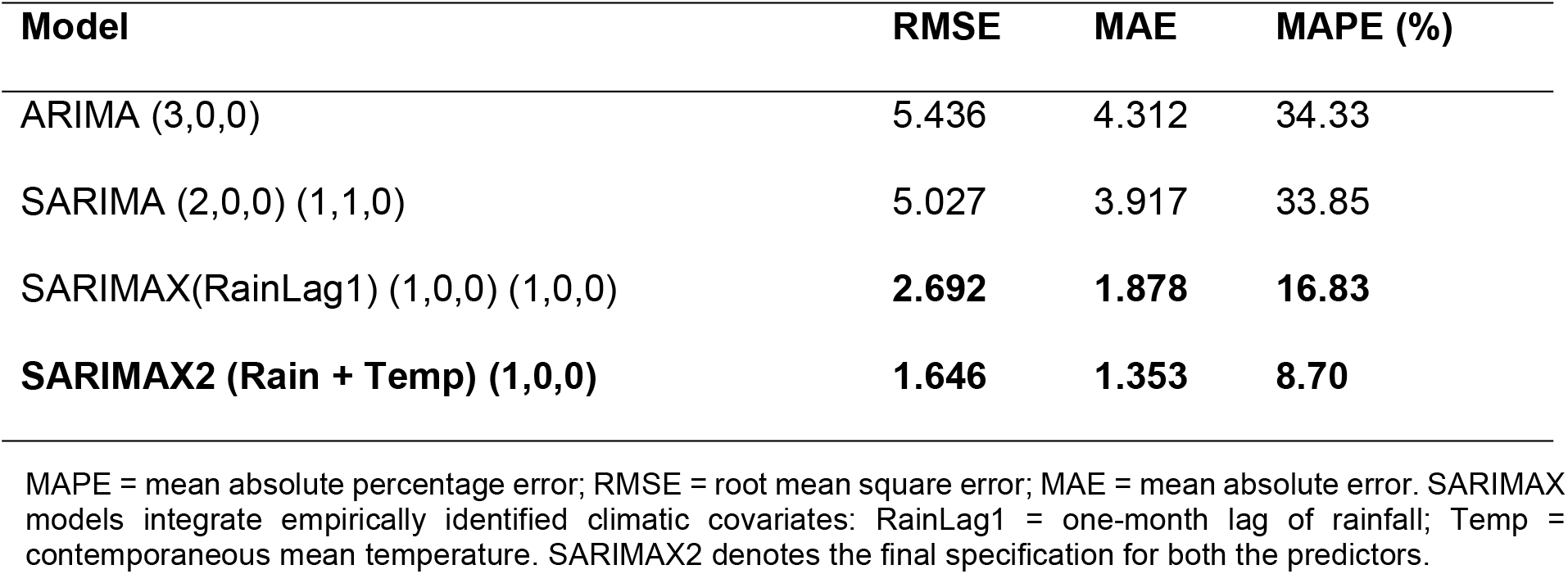
Out-of-Sample Forecasting Performance of candidate models.

| Model | RMSE | MAE | MAPE (%) |
| --- | --- | --- | --- |
| ARIMA (3,0,0) | 5.436 | 4.312 | 34.33 |
| SARIMA (2,0,0) (1,1,0) | 5.027 | 3.917 | 33.85 |
| SARIMAX(RainLag1) (1,0,0) (1,0,0) | **2.692** | **1.878** | **16.83** |
| **SARIMAX2 (Rain + Temp) (1,0,0)** | **1.646** | **1.353** | **8.70** |

MAPE = mean absolute percentage error; RMSE = root mean square error; MAE = mean absolute error. SARIMAX models integrate empirically identified climatic covariates: RainLag1 = one-month lag of rainfall; Temp = contemporaneous mean temperature. SARIMAX2 denotes the final specification for both the predictors.

### Model Performance and selection

Three classes of time series models were evaluated: ARIMA, SARIMA, and SARIMAX.

The incorporation of exogenous climatic variables in the SARIMAX model improved the model fit and forecasting performance compared to the univariate ARIMA and SARIMA models.

Performance metrics are presented in Table 2.

The performance metrics showed improved predictive accuracy in the SARIMAX model, with lower RMSE and MAE values and a Mean Absolute Percentage Error (MAPE) of 8.7% (Table 2).

The observed and fitted values demonstrated closer agreement for the final SARIMAX model than for the ARIMA and SARIMA models (Figure 4).

**Figure 4:**
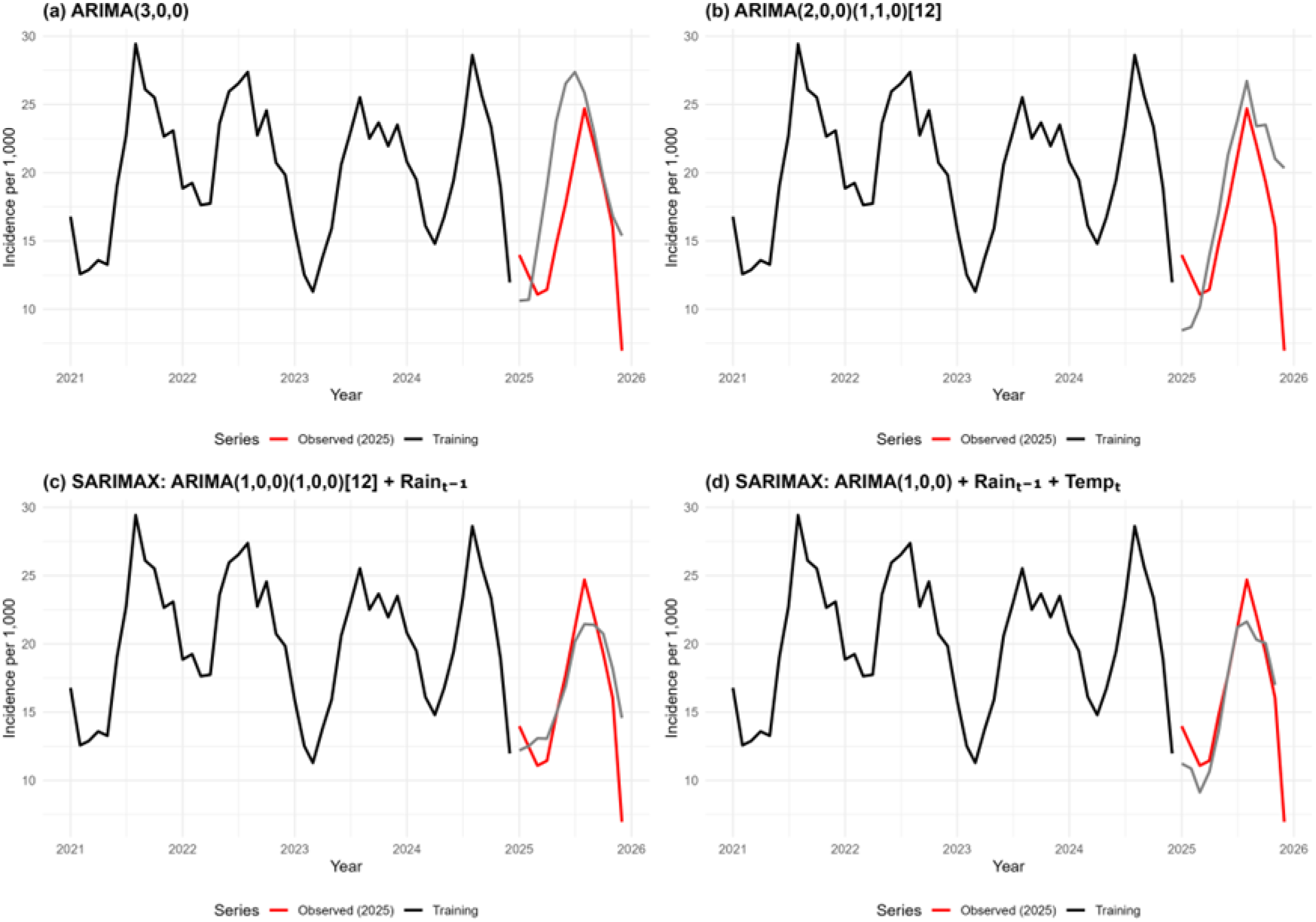
Out-of-Sample Forecasting Performance Comparison of ARIMA, SARIMA and SARIMAX models. Observed versus predicted malaria incidence values using the 2025 test dataset.

### Final SARIMAX Model Parameters

The final SARIMAX model demonstrated strong autoregressive structure and improved predictive performance.

Model coefficients are presented in Table 3.

**Table 3II:** Final SARIMAX model parameter estimates.

| Coefficient | Estimate | Std. Error | z-value | p-value |
| --- | --- | --- | --- | --- |
| ar1 | 0.8819 | 0.1250 | 7.0561 | <0.001 |
| ar2 | 0.0727 | 0.1698 | 0.4281 | 0.669 |
| ar3 | -0.4180 | 0.1281 | -3.2621 | 0.0011 |
| intercept | 39.2867 | 9.2001 | 4.2703 | <0.001 |
| rain_l1 | 0.0008 | 0.0030 | 0.2671 | 0.7894 |
| temp0 | -0.7445 | 0.3440 | -2.1642 | 0.0304 |

AR = autoregressive term; Std = standard deviation Error = standard error of the estimate. Rainfall was measured in millimetres (lagged by one month) and temperature in degrees Celsius (contemporaneous). Positive coefficients indicate a direct relationship with malaria incidence, while negative coefficients indicate an inverse relationship.

### Forecasting malaria incidence for 2026

The final SARIMAX model was used to generate out-of-sample forecasts for the year 2026. The forecast results indicated sustained malaria transmission throughout the year, with a consistent seasonal peak occurring between June and August (Figure 5).

**Figure 5:**
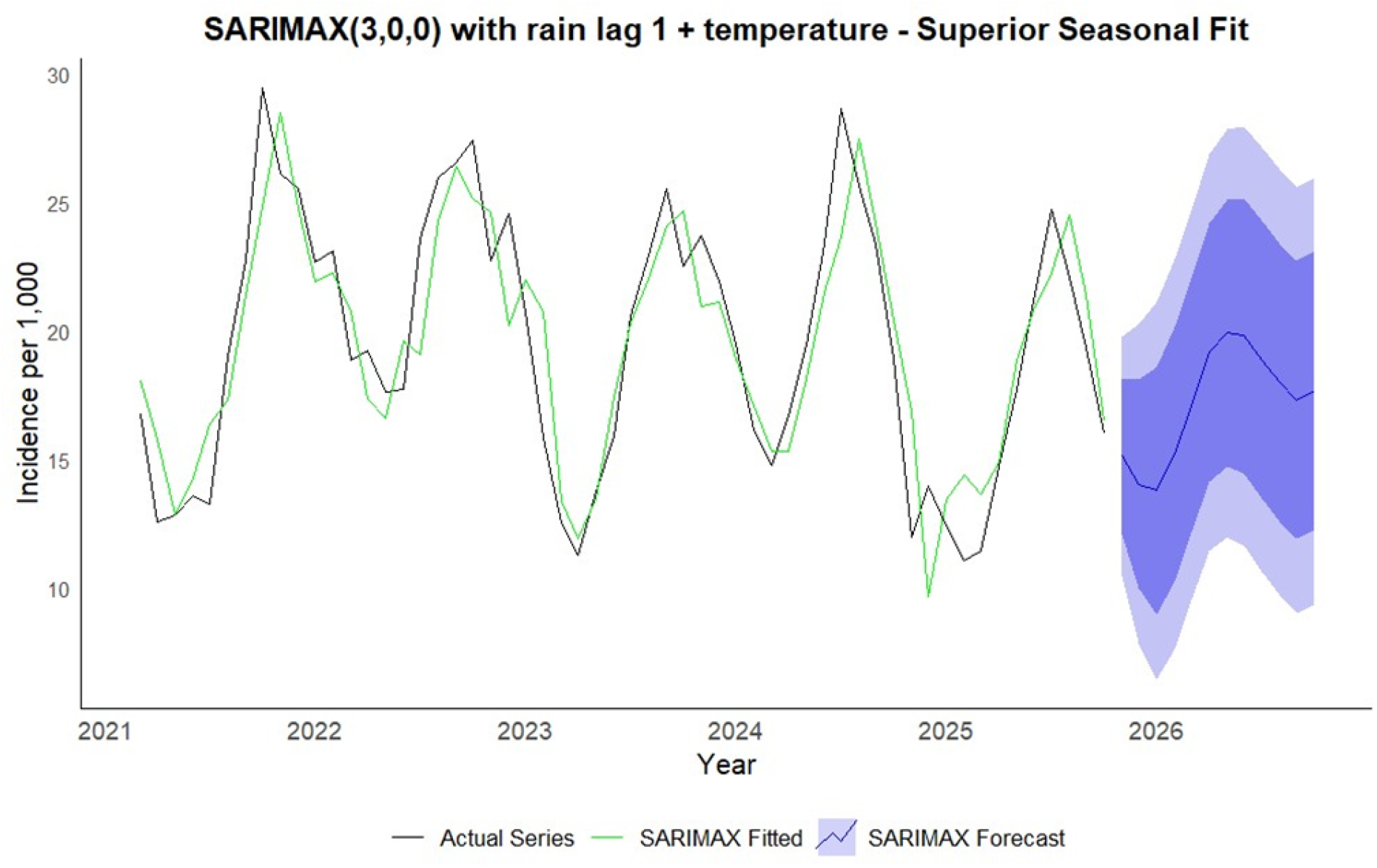
SARIMAX forecast of malaria incidence in Nasarawa State. Forecast monthly malaria incidence with 95% confidence intervals generated from the final SARIMA model incorporating lagged rainfall and contemporaneous temperature.

Monthly forecast values are presented in Table 4.

**Table 4.** Monthly forecasted malaria incidence per 1,000 population for 2026.

| Month 2026 | Mean | 95% LCL | 95% UCL |
| --- | --- | --- | --- |
| Jan | 15.15 | 10.51 | 19.78 |
| Feb | 14.06 | 7.88 | 20.24 |
| Mar | 13.81 | 6.48 | 21.14 |
| Apr | 15.20 | 7.64 | 22.76 |
| May | 17.09 | 9.53 | 24.65 |
| Jun | 19.16 | 11.49 | 26.84 |
| Jul | 19.93 | 12.01 | 27.85 |
| Aug | 19.81 | 11.67 | 27.96 |
| Sep | 18.88 | 10.64 | 27.11 |
| Oct | 18.00 | 9.76 | 26.25 |
| Nov | 17.32 | 9.06 | 25.57 |
| Dec | 17.67 | 9.38 | 25.96 |

\*Forecasted incidence values are accompanied by 95% confidence intervals calculated as $\hat{Y}_t \pm 1.96 \times S$ , where $\hat{Y}_t$ is the forecasted incidence and $S=2.366S$ is the innovation standard error of the fitted SARIMAX model. The negative lower bounds are truncated to zero for interpretability.

## DISCUSSION

This study demonstrated that malaria incidence in Nasarawa State follows a pronounced seasonal pattern, strongly influenced by climatic variability, particularly rainfall and temperature. The observed seasonal transmission peaks occurring between June and August correspond closely with the rainy season and reinforce established eco-epidemiological theories linking environmental conditions with vector ecology, parasite development, and malaria transmission dynamics [2,3]. Similar seasonal malaria patterns have been reported across the malaria-endemic regions of sub-Saharan Africa, where rainfall and temperature remain major determinants of vector abundance and transmission intensity [4,5].

The strong seasonal component identified in this study reflects the ecological dependence of *Anopheles* mosquito breeding on rainfall availability. Increased rainfall contributes to the formation and persistence of stagnant water collections, which serve as breeding habitats for mosquito vectors. However, studies on the effects of rainfall on malaria transmission are rare. The one-month lag association observed between rainfall and malaria incidence is biologically plausible and consistent with the developmental cycle of *Anopheles* mosquitoes and the extrinsic incubation period of *Plasmodium* parasites within the vector. Following rainfall events, a period of time is generally required for larval maturation, increased adult mosquito density, successful blood feeding, parasite development, and eventual transmission to human hosts. Similar lag structures have been documented in malaria forecasting studies conducted in Nigeria, Ethiopia, Sierra Leone, and other malaria-endemic African settings [5,8,9].

The inverse relationship observed between temperature and malaria incidence may reflect the complex nonlinear relationship between temperature and malaria transmission dynamics. While moderate increases in temperature may accelerate parasite development and mosquito biting activity, excessively high temperatures may adversely affect mosquito survival, feeding behaviour, and vector competence. Previous thermal biology studies have demonstrated that malaria transmission efficiency declines when environmental temperatures exceed optimal physiological thresholds for mosquito survival and parasite development [3]. Given the relatively high ambient temperatures characteristic of North-Central Nigeria, elevated temperatures during certain periods may therefore reduce transmission efficiency despite favourable rainfall conditions.

These findings also highlight the importance of incorporating climatic variables into malaria forecasting systems. The SARIMAX model incorporating rainfall and temperature outperformed the univariate ARIMA and SARIMA models across multiple forecasting performance metrics, including the RMSE, MAE, and MAPE. This finding suggests that malaria transmission dynamics in Nasarawa State are not solely dependent on historical temporal trends but are also strongly influenced by contemporaneous and lagged environmental conditions. By integrating climatic predictors, the SARIMAX model was better able to capture both seasonal autocorrelation and climate-driven transmission variability, thereby improving the predictive performance.

The superior performance of the climate-informed forecasting models observed in this study was consistent with the findings of previous malaria forecasting studies. Bakare et al. demonstrated the utility of SARIMA-based approaches for modelling malaria trends in selected Nigerian states [7]. Similarly, a recent study conducted in Sierra Leone reported improved malaria forecasting accuracy when climatic variability was incorporated into the SARIMA and SARIMAX frameworks [8]. In Ethiopia, Gelaw and Abera demonstrated that integrating rainfall and temperature as exogenous regressors significantly improves malaria forecasting performance within a SARIMAX framework in a resource-limited urban setting [9]. Comparable findings have also been reported in South Africa, where SARIMA-based models were successfully applied to support malaria surveillance and elimination planning in the Limpopo Province [10].

In addition to conventional statistical forecasting models, recent advances in machine learning and artificial intelligence have expanded the opportunities for climate-informed malaria prediction. Transformer-based deep-learning models using high-resolution environmental datasets have recently demonstrated promising predictive performance for malaria forecasting [11]. Generalized linear modelling approaches have also been successfully applied in the malaria-endemic regions of Senegal [12]. Nevertheless, despite these methodological advances, the SARIMA and SARIMAX models remain highly valuable for operational public health use in low-resource settings because they are transparent, interpretable, computationally efficient, and compatible with the routinely available surveillance data.

One of the major strengths of this study was the use of routinely available surveillance and meteorological datasets, which demonstrated the feasibility of integrating predictive analytics into existing malaria surveillance systems. The use of DHIS2-derived malaria surveillance data further highlights the operational value of routinely collected health information in supporting evidence-based malaria control planning and preparedness in resource-limited settings. Because the modelling framework utilizes routinely collected monthly data, the approach may be adaptable for implementation within subnational malaria surveillance programs and climate-informed early warning systems in similar endemic settings.

The study additionally contributes to the growing body of evidence supporting climate-sensitive malaria surveillance in Nigeria, where predictive modelling remains relatively underutilized despite substantial malaria burden and increasing climate variability. Integrating forecasting models into malaria surveillance systems may support more proactive allocation of malaria commodities, targeted vector control interventions, seasonal preparedness planning, and optimization of limited public health resources during high transmission periods. Despite these strengths, several limitations should be acknowledged. First, the ecological study design limits causal inference at the individual level because aggregated population-level data were analysed. Second, routine surveillance datasets may be affected by under-reporting, reporting delays, incomplete documentation, diagnostic variability across facilities, and inconsistencies in health facility reporting practices. Third, several important determinants of malaria transmission were not included in the present model, including humidity, vegetation indices, land use patterns, socioeconomic factors, population movement, vector control coverage, and entomological variables, all of which may further influence transmission dynamics and forecasting accuracy [13]. Additionally, the relatively short duration of available time-series data may limit the long-term forecasting stability and external generalizability.

Nevertheless, the findings provide important evidence supporting climate-informed malaria forecasting in Nigeria and contribute to the growing body of literature supporting the use of time-series modelling approaches for malaria surveillance and preparedness.

## CONCLUSION

Malaria incidence in Nasarawa State demonstrates pronounced seasonal variation, which is strongly influenced by rainfall and temperature. The study identified clear seasonal transmission peaks during the rainy season, with rainfall exhibiting a delayed positive association with malaria incidence. Incorporating climatic variables into the SARIMAX framework substantially improved forecasting performance compared with conventional ARIMA and SARIMA models, highlighting the importance of climate-sensitive modelling approaches for malaria prediction.

These findings demonstrate the operational potential of integrating routine surveillance and meteorological data for climate-informed malaria forecasting in endemic settings. Such forecasting approaches may support more proactive malaria preparedness, improved timing of vector control interventions, optimization of resource allocation, and strengthening of malaria early warning systems within resource-limited health systems.

Although the present model demonstrated strong predictive performance, future studies incorporating additional environmental, socioeconomic, entomological, and intervention-related variables across broader geographic settings may further improve the forecasting accuracy and external applicability. The continued integration of predictive analytics into routine malaria surveillance systems may contribute substantially to evidence-based malaria control planning and climate-resilient public health preparedness in Nigeria and other malaria-endemic regions.

## AUTHOR CONTRIBUTIONS

Conceptualization: Grace Iberi Iheanacho

Data curation: Grace Iberi Iheanacho

Formal analysis: Grace Iberi Iheanacho

Methodology: Grace Iberi Iheanacho, Maxwell Azubike Ijomah

Supervision: Maxwell Azubike Ijomah, Ibidabo David Alabere

Writing – original draft: Grace Iberi Iheanacho

Writing – review & editing: Maxwell Azubike Ijomah, Ibidabo David Alabere

## DATA AVAILABILITY STATEMENT

The aggregated malaria surveillance dataset analysed during the current study was derived from the District Health Information Software 2 (DHIS2) platform of the Nasarawa State Ministry of Health under administrative approval. Due to institutional data governance restrictions, raw facility-level data are not publicly available. However, the minimal anonymized analytical dataset and modelling scripts supporting the findings of this study may be made available from the corresponding author upon reasonable request and with permission from the Nasarawa State Ministry of Health. Meteorological data were obtained from the Nigerian Meteorological Agency (NiMet).

## ACKNOWLEDGEMENTS

The authors acknowledge the Corona Management Systems, Nasarawa State Ministry of Health, DHIS2 team, Nigerian Meteorological Agency (NiMet), and ACE-PUTOR for their support and data access.

